## Supplementary table 2 for "Psychometric properties of upper limb kinematics during functional tasks in children and adolescents with dyskinetic cerebral palsy"

| Table S2: Intra-class correlation coefficients (ICC) and standard error of measurement (SEM) and change in ICC and SEM values with each number of different repetitions, expressed in %. | | | | | | | | | | | | | | | | | | | | | | | | | | | | | | | | | | | | | | |
| --- | --- | --- | --- | --- | --- | --- | --- | --- | --- | --- | --- | --- | --- | --- | --- | --- | --- | --- | --- | --- | --- | --- | --- | --- | --- | --- | --- | --- | --- | --- | --- | --- | --- | --- | --- | --- | --- | --- |
| REACH FORWARD | | | | | | | | | | | | | | | | | | | | | | | | | | | | | | | | | | | | | | |
|  | Elevation  Plane | | Shoulder  Elevation | | | Shoulder  Rotation | | | Elbow  Fl/Ext | | | Elbow  Pro/Sup | | | Wrist  Fl/Ext | | | Wrist  Deviation | | | Scapular  Pro/Retr | | | Scapular  Rotation | | | Scapular  Tilting | | | Trunk  Fl/Ext | | | Trunk  Lateral Fl | | | Trunk  Axial Rot | | |
|  | ICC | SEM | | ICC | SEM | | ICC | SEM | | ICC | SEM | | ICC | SEM | | ICC | SEM | | ICC | SEM | | ICC | SEM | | ICC | SEM | | ICC | SEM | | ICC | SEM | | ICC | SEM | | ICC | SEM |
| TD |  |  | |  |  | |  |  | |  |  | |  |  | |  |  | |  |  | |  |  | |  |  | |  |  | |  |  | |  |  | |  |  |
| 2 REP | 0.88 | 2.82 | | 0.94 | 2.12 | | 0.98 | 11.61 | | 0.84 | 4.36 | | 0.91 | 6.65 | | 0.86 | 4.58 | | 0.72 | 3.60 | | 0.98 | 1.39 | | 0.94 | 3.44 | | 0.95 | 2.75 | | 0.94 | 1.30 | | 0.86 | 2.09 | | 0.80 | 2.42 |
| 4 REP | 0.88 | 2.73 | | 0.96 | 1.91 | | 0.97 | 9.74 | | 0.86 | 4.09 | | 0.94 | 5.46 | | 0.89 | 4.02 | | 0.53 | 4.35 | | 0.97 | 1.97 | | 0.96 | 2.83 | | 0.96 | 2.33 | | 0.81 | 2.17 | | 0.85 | 2.34 | | 0.86 | 2.07 |
| 6 REP | 0.87 | 2.76 | | 0.94 | 2.13 | | 0.97 | 9.70 | | 0.87 | 4.13 | | 0.93 | 6.18 | | 0.89 | 3.99 | | 0.55 | 4.16 | | 0.97 | 1.91 | | 0.97 | 2.59 | | 0.97 | 2.29 | | 0.85 | 2.00 | | 0.82 | 2.49 | | 0.84 | 2.28 |
| 8 REP | 0.87 | 2.82 | | 0.93 | 2.24 | | 0.97 | 9.66 | | 0.86 | 4.19 | | 0.93 | 6.20 | | 0.90 | 3.87 | | 0.61 | 3.86 | | 0.97 | 1.91 | | 0.97 | 2.45 | | 0.97 | 2.16 | | 0.85 | 2.00 | | 0.82 | 2.43 | | 0.83 | 2.27 |
| 10 REP | 0.87 | 2.76 | | 0.94 | 2.20 | | 0.97 | 9.18 | | 0.86 | 4.29 | | 0.92 | 6.65 | | 0.90 | 3.98 | | 0.62 | 3.76 | | 0.97 | 1.82 | | 0.97 | 2.45 | | 0.96 | 2.34 | | 0.86 | 1.95 | | 0.80 | 2.44 | | 0.83 | 2.27 |
| % 2-4 | -0.16 | -3.30 | | 1.2 | -9.27 | | -1.7 | -16.1 | | 1.8 | -6.04 | | 2.7 | -17.8 | | 2.4 | -12.1 | | -27.0 | 17.2 | | -1.5 | 29.8 | | 2.1 | -17.9 | | 1.4 | -15.5 | | -13.3 | 40.02 | | -1.3 | 10.0 | | 7.5 | -14.2 |
| % 4-6 | -1.1 | 1.2 | | -1.5 | 9.8 | | 0.05 | -0.36 | | 1.2 | 0.94 | | -1.2 | 10.8 | | 0.87 | -0.68 | | 2.8 | -4.2 | | 0.30 | -3.5 | | 0.64 | -6.7 | | 0.11 | -1.38 | | 3.7 | -7.9 | | -4.2 | 6.1 | | -3.2 | 8.6 |
| % 6-8 | -0.36 | 2.0 | | -0.83 | 5.0 | | 0.35 | -0.33 | | -1.2 | 1.2 | | 0.54 | 0.29 | | 1.1 | -2.6 | | 8.1 | -6.9 | | -0.03 | 0.12 | | 0.27 | -3.8 | | 0.37 | -4.6 | | 0.01 | -0.17 | | -0.08 | -2.2 | | -0.38 | -0.32 |
| % 8-10 | 0.90 | -1.9 | | 0.24 | -2.0 | | 0.22 | -4.2 | | -0.31 | 2.30 | | -0.92 | 6.8 | | -0.36 | 2.4 | | 1.9 | -2.4 | | 0.26 | -4.3 | | -0.05 | -0.05 | | -0.56 | 6.5 | | 1.2 | -2.2 | | -1.5 | 0.19 | | -0.33 | -0.04 |
| DCP |  |  | |  |  | |  |  | |  |  | |  |  | |  |  | |  |  | |  |  | |  |  | |  |  | |  |  | |  |  | |  |  |
| 2 REP | 0.89 | 5.31 | | 0.92 | 3.45 | | 0.91 | 9.67 | | 0.90 | 5.39 | | 0.93 | 5.42 | | 0.72 | 11.2 | | 0.86 | 6.28 | | 0.95 | 2.97 | | 0.97 | 3.23 | | 0.98 | 1.90 | | 0.89 | 3.97 | | 0.34 | 6.38 | | 0.83 | 4.55 |
| 4 REP | 0.90 | 4.83 | | 0.88 | 4.28 | | 0.91 | 9.12 | | 0.74 | 8.96 | | 0.93 | 5.19 | | 0.64 | 13.4 | | 0.66 | 8.59 | | 0.93 | 3.31 | | 0.95 | 3.78 | | 0.94 | 3.64 | | 0.84 | 4.62 | | 0.55 | 5.18 | | 0.80 | 4.40 |
| 6 REP | 0.90 | 5.06 | | 0.86 | 4.55 | | 0.92 | 8.91 | | 0.75 | 9.19 | | 0.91 | 5.89 | | 0.69 | 12.9 | | 0.69 | 7.92 | | 0.92 | 3.45 | | 0.95 | 3.60 | | 0.93 | 3.82 | | 0.83 | 4.59 | | 0.59 | 4.99 | | 0.79 | 5.13 |
| 8 REP | 0.87 | 5.83 | | 0.87 | 4.38 | | 0.93 | 8.63 | | 0.78 | 8.66 | | 0.91 | 5.96 | | 0.69 | 12.8 | | 0.74 | 7.75 | | 0.92 | 3.46 | | 0.96 | 3.41 | | 0.94 | 3.47 | | 0.85 | 4.34 | | 0.63 | 4.94 | | 0.75 | 5.31 |
| 10 REP | 0.87 | 5.82 | | 0.86 | 4.57 | | 0.94 | 8.59 | | 0.79 | 8.34 | | 0.91 | 5.83 | | 0.70 | 13.0 | | 0.76 | 7.51 | | 0.92 | 3.40 | | 0.96 | 3.46 | | 0.94 | 3.45 | | 0.85 | 4.36 | | 0.65 | 4.81 | | 0.77 | 5.13 |
| % 2-4 | 1.7 | -8.3 | | -4.9 | 18.2 | | 0.11 | -5.7 | | -18.2 | 38.8 | | 0.32 | -3.76 | | -11.6 | 16.5 | | -23.1 | 26.9 | | -1.5 | 9.76 | | -1.9 | 14.6 | | -4.7 | 45.43 | | -5.6 | 14.09 | | 32.7 | -18.8 | | -3.1 | -2.98 |
| % 4-6 | -0.21 | 4.02 | | -1.9 | 5.88 | | 0.71 | -2.1 | | 1.5 | 2.50 | | -2.3 | 11.6 | | 6.6 | -3.6 | | 3.8 | -7.9 | | -0.97 | 4.17 | | 0.69 | -4.9 | | -0.48 | 4.79 | | -0.69 | -0.69 | | 6.4 | -3.1 | | -1.2 | 13.8 |
| % 6-8 | -4.0 | 13.1 | | 0.99 | -3.58 | | 0.92 | -3.0 | | 2.9 | -5.79 | | 0.00 | 1.23 | | 0.81 | -0.92 | | 5.2 | -1.95 | | -0.48 | 0.09 | | 0.40 | -4.90 | | 1.0 | -9.26 | | 2.2 | -5.27 | | 5.7 | -0.78 | | -4.3 | 3.39 |
| % 8-10 | 0.21 | -0.17 | | -1.1 | 4.07 | | 0.86 | -0.41 | | 2.0 | -3.45 | | 0.61 | -2.13 | | 1.3 | 1.23 | | 3.1 | -2.83 | | 0.39 | -1.75 | | -0.05 | 1.24 | | -0.08 | -0.59 | | 0.23 | 0.41 | | 2.4 | -1.93 | | 1.9 | -3.33 |
| TD = typically developing participants; DCP = participants with dyskinetic cerebral palsy. Fl = flexion; Ext = extension; Pro = protraction; Retr = retraction; Rot = rotation, REP = repetitions. | | | | | | | | | | | | | | | | | | | | | | | | | | | | | | | | | | | | | | |

| Table S2 (continued) | | | | | | | | | | | | | | | | | | | | | | | | | | |
| --- | --- | --- | --- | --- | --- | --- | --- | --- | --- | --- | --- | --- | --- | --- | --- | --- | --- | --- | --- | --- | --- | --- | --- | --- | --- | --- |
| REACH AND GRASP VERTICAL | | | | | | | | | | | | | | | | | | | | | | | | | | |
|  | Elevation  Plane | | Shoulder  Elevation | | Shoulder  Rotation | | Elbow  Fl/Ext | | Elbow  Pro/Sup | | Wrist  Fl/Ext | | Wrist  Deviation | | Scapular  Pro/Retr | | Scapular  Rotation | | Scapular  Tilting | | Trunk  Fl/Ext | | Trunk  Lateral Fl | | Trunk  Axial Rot | |
|  | ICC | SEM | ICC | SEM | ICC | SEM | ICC | SEM | ICC | SEM | ICC | SEM | ICC | SEM | ICC | SEM | ICC | SEM | ICC | SEM | ICC | SEM | ICC | SEM | ICC | SEM |
| TD |  |  |  |  |  |  |  |  |  |  |  |  |  |  |  |  |  |  |  |  |  |  |  |  |  |  |
| 2 REP | 0.89 | 3.15 | 0.91 | 2.47 | 1.00 | 3.56 | 0.87 | 5.70 | 0.88 | 4.26 | 0.93 | 6.41 | 0.94 | 4.86 | 0.92 | 1.60 | 0.94 | 3.99 | 0.96 | 1.94 | 0.92 | 2.33 | 0.83 | 3.78 | 0.45 | 2.95 |
| 4 REP | 0.83 | 3.31 | 0.90 | 2.79 | 1.00 | 3.14 | 0.81 | 4.69 | 0.91 | 5.06 | 0.86 | 6.62 | 0.81 | 4.11 | 0.90 | 1.75 | 0.94 | 3.19 | 0.95 | 1.81 | 0.85 | 2.12 | 0.76 | 2.83 | 0.57 | 2.92 |
| 6 REP | 0.83 | 3.16 | 0.90 | 2.97 | 1.00 | 2.88 | 0.83 | 4.93 | 0.92 | 4.50 | 0.81 | 6.56 | 0.84 | 4.02 | 0.88 | 1.93 | 0.95 | 2.95 | 0.96 | 1.76 | 0.85 | 2.38 | 0.73 | 2.87 | 0.59 | 3.21 |
| 8 REP | 0.85 | 3.34 | 0.91 | 2.78 | 1.00 | 2.79 | 0.82 | 5.72 | 0.93 | 4.33 | 0.82 | 6.85 | 0.85 | 4.16 | 0.89 | 2.17 | 0.95 | 3.24 | 0.95 | 1.79 | 0.86 | 2.24 | 0.74 | 2.98 | 0.56 | 3.03 |
| 10 REP | 0.86 | 3.25 | 0.91 | 2.69 | 1.00 | 2.87 | 0.83 | 5.66 | 0.93 | 4.51 | 0.83 | 6.84 | 0.85 | 4.08 | 0.88 | 2.09 | 0.95 | 3.22 | 0.95 | 1.90 | 0.87 | 2.16 | 0.76 | 2.79 | 0.59 | 3.01 |
| % 2-4 | -6.0 | 4.8 | -0.77 | 10.6 | 0.03 | -11.7 | -6.4 | -17.7 | 3.2 | 15.7 | -7.1 | 3.0 | -13.7 | -15.6 | -2.3 | 7.0 | -0.36 | -20.1 | -0.33 | -6.6 | -7.0 | -8.9 | -8.8 | -25.3 | 20.6 | -0.84 |
| % 4-6 | 0.01 | -4.5 | 0.39 | 6.1 | -0.03 | -7.3 | 2.1 | 4.2 | 0.30 | -11.0 | -4.9 | -0.79 | 3.0 | -1.8 | -2.1 | 7.9 | 1.1 | -6.1 | 0.67 | -2.5 | -0.15 | 10.9 | -3.2 | 1.3 | 2.8 | 9.0 |
| % 6-8 | 1.6 | 5.6 | 0.95 | -6.3 | 0.00 | -2.7 | -1.4 | 13.9 | 1.2 | -3.4 | 1.1 | 4.2 | 1.3 | 2.9 | 0.60 | 11.1 | 0.34 | 7.4 | -1.6 | 1.7 | 0.96 | -5.6 | 1.8 | 2.8 | -5.3 | -5.4 |
| % 8-10 | 1.2 | -2.7 | 0.00 | -3.2 | -0.02 | 2.4 | 0.99 | -1.1 | 0.22 | 3.6 | 0.9 | -0.11 | -0.31 | -1.5 | -0.52 | -3.5 | -0.27 | -0.66 | 0.16 | 5.4 | 0.73 | -3.3 | 2.1 | -5.1 | 5.5 | -0.64 |
| DCP |  |  |  |  |  |  |  |  |  |  |  |  |  |  |  |  |  |  |  |  |  |  |  |  |  |  |
| 2 REP | 0.89 | 5.03 | 0.90 | 3.59 | 0.95 | 4.53 | 0.65 | 6.30 | 0.97 | 9.82 | 0.81 | 7.02 | 0.73 | 3.11 | 0.98 | 2.81 | 0.89 | 4.36 | 0.97 | 3.32 | 0.85 | 3.26 | 0.52 | 2.92 | 0.78 | 7.79 |
| 4 REP | 0.87 | 5.96 | 0.88 | 3.84 | 0.96 | 4.31 | 0.80 | 7.68 | 0.96 | 8.10 | 0.78 | 9.98 | 0.82 | 7.14 | 0.97 | 3.05 | 0.93 | 4.48 | 0.98 | 3.24 | 0.87 | 4.00 | 0.64 | 3.56 | 0.77 | 7.21 |
| 6 REP | 0.88 | 5.95 | 0.87 | 3.84 | 0.97 | 4.52 | 0.76 | 7.33 | 0.97 | 7.69 | 0.79 | 11.1 | 0.83 | 6.72 | 0.96 | 3.35 | 0.95 | 4.18 | 0.98 | 3.09 | 0.85 | 4.03 | 0.64 | 3.79 | 0.72 | 7.02 |
| 8 REP | 0.86 | 5.67 | 0.89 | 3.62 | 0.97 | 4.52 | 0.71 | 7.47 | 0.97 | 7.20 | 0.79 | 10.7 | 0.82 | 6.39 | 0.95 | 3.25 | 0.93 | 4.08 | 0.98 | 3.62 | 0.87 | 3.94 | 0.65 | 3.82 | 0.75 | 7.14 |
| 10 REP | 0.87 | 5.45 | 0.90 | 3.64 | 0.97 | 4.68 | 0.72 | 7.20 | 0.97 | 7.00 | 0.80 | 10.6 | 0.82 | 6.53 | 0.96 | 3.39 | 0.93 | 4.12 | 0.97 | 3.57 | 0.88 | 3.82 | 0.69 | 3.67 | 0.76 | 7.05 |
| % 2-4 | -2.1 | 15.6 | -2.1 | 6.6 | 1.4 | -4.9 | 18.3 | 18.0 | -1.2 | -17.5 | -3.1 | 26.6 | 11.3 | 56.4 | -0.68 | 6.9 | 4.4 | 2.8 | 0.48 | -2.0 | 2.4 | 18.4 | 18.4 | 16.9 | -0.99 | -7.5 |
| % 4-6 | 0.84 | -0.14 | -1.4 | 0.08 | 0.61 | 4.6 | -4.3 | -4.6 | 0.98 | -4.2 | 1.2 | 10.3 | 1.3 | -5.8 | -0.73 | 9.0 | 1.2 | -6.7 | 0.09 | -4.3 | -2.8 | 0.64 | 0.43 | 6.1 | -5.7 | -2.4 |
| % 6-8 | -1.2 | -4.7 | 2.0 | -5.7 | 0.24 | -0.17 | -6.2 | 1.9 | 0.29 | -5.0 | -0.75 | -4.2 | -1.5 | -4.7 | -1.0 | -2.9 | -1.5 | -2.4 | -0.13 | 14.7 | 2.8 | -2.1 | 0.84 | 0.59 | 2.9 | 1.5 |
| % 8-10 | 1.1 | -3.8 | 1.0 | 0.42 | -0.17 | 3.6 | 1.2 | -3.6 | -0.16 | -2.0 | 1.5 | -0.70 | 0.15 | 2.0 | 0.39 | 4.1 | 0.00 | 0.96 | -0.28 | -1.4 | 0.90 | -3.04 | 5.1 | -3.7 | 1.4 | -1.17 |
| TD = typically developing participants; DCP = participants with dyskinetic cerebral palsy. Fl = flexion; Ext = extension; Pro = protraction; Retr = retraction; Rot = rotation, REP = repetitions. | | | | | | | | | | | | | | | | | | | | | | | | | | |

| Table S2 (continued) | | | | | | | | | | | | | | | | | | | | | | | | | | | | | | | | | |
| --- | --- | --- | --- | --- | --- | --- | --- | --- | --- | --- | --- | --- | --- | --- | --- | --- | --- | --- | --- | --- | --- | --- | --- | --- | --- | --- | --- | --- | --- | --- | --- | --- | --- |
| REACH SIDEWAYS | | | | | | | | | | | | | | | | | | | | | | | | | | | | | | | | | |
|  | Elevation  Plane | | Shoulder  Elevation | | Shoulder  Rotation | | Elbow  Fl/Ext | | Elbow  Pro/Sup | | Wrist  Fl/Ext | | Wrist  Deviation | | | Scapular  Pro/Retr | | | Scapular  Rotation | | | Scapular  Tilting | | | Trunk  Fl/Ext | | | Trunk  Lateral Fl | | | Trunk  Axial Rot | | |
|  | ICC | SEM | ICC | SEM | ICC | SEM | ICC | SEM | ICC | SEM | ICC | SEM | | ICC | SEM | | ICC | SEM | | ICC | SEM | | ICC | SEM | | ICC | SEM | | ICC | SEM | | ICC | SEM |
| TD |  |  |  |  |  |  |  |  |  |  |  |  | |  |  | |  |  | |  |  | |  |  | |  |  | |  |  | |  |  |
| 2 REP | 0.92 | 3.19 | 0.89 | 3.23 | 0.98 | 2.77 | 0.86 | 6.30 | 0.97 | 4.41 | 0.86 | 5.41 | | 0.54 | 4.66 | | 0.98 | 2.29 | | 0.96 | 2.37 | | 0.97 | 2.21 | | 0.97 | 1.13 | | 0.84 | 2.15 | | 0.74 | 2.83 |
| 4 REP | 0.88 | 4.00 | 0.90 | 3.12 | 0.97 | 3.45 | 0.73 | 7.87 | 0.97 | 4.30 | 0.91 | 4.37 | | 0.68 | 4.00 | | 0.98 | 2.48 | | 0.96 | 2.25 | | 0.97 | 2.13 | | 0.94 | 1.74 | | 0.81 | 2.22 | | 0.79 | 2.60 |
| 6 REP | 0.88 | 3.98 | 0.91 | 2.98 | 0.97 | 3.33 | 0.77 | 7.38 | 0.97 | 4.23 | 0.91 | 4.30 | | 0.70 | 3.78 | | 0.98 | 2.45 | | 0.97 | 2.18 | | 0.97 | 2.18 | | 0.94 | 1.69 | | 0.83 | 2.16 | | 0.75 | 2.73 |
| 8 REP | 0.87 | 4.14 | 0.92 | 2.90 | 0.97 | 3.40 | 0.81 | 6.85 | 0.97 | 4.39 | 0.91 | 4.25 | | 0.70 | 3.84 | | 0.98 | 2.42 | | 0.96 | 2.30 | | 0.97 | 2.15 | | 0.93 | 1.76 | | 0.81 | 2.26 | | 0.76 | 2.72 |
| 10 REP | 0.86 | 4.17 | 0.92 | 2.90 | 0.96 | 3.64 | 0.81 | 6.72 | 0.97 | 4.46 | 0.90 | 4.52 | | 0.71 | 3.79 | | 0.98 | 2.47 | | 0.97 | 2.24 | | 0.97 | 2.25 | | 0.93 | 1.85 | | 0.80 | 2.33 | | 0.76 | 2.71 |
| % 2-4 | -4.4 | 19.3 | 1.4 | -3.4 | -1.1 | 18.6 | -15.1 | 20.0 | 0.24 | -2.6 | 5.8 | -19.4 | | 19.0 | -14.2 | | -0.6 | 7.7 | | 0.49 | -5.1 | | 0.20 | -3.4 | | -3.9 | 32.7 | | -3.0 | 2.8 | | 6.2 | -8.1 |
| % 4-6 | -0.68 | -0.48 | 0.91 | -4.5 | 0.21 | -3.1 | 4.4 | -6.2 | -0.01 | -1.5 | -0.16 | -1.3 | | 2.8 | -4.7 | | 0.00 | -1.1 | | 0.44 | -3.1 | | -0.15 | 2.3 | | 0.51 | -2.5 | | 2.0 | -2.3 | | -4.1 | 4.7 |
| % 6-8 | -1.1 | 3.9 | 0.63 | -2.4 | -0.17 | 1.74 | 4.3 | -6.8 | -0.26 | 3.5 | -0.01 | -0.79 | | -0.33 | 1.30 | | 0.06 | -1.4 | | -0.37 | 5.0 | | 0.10 | -1.7 | | -0.74 | 3.8 | | -1.8 | 4.1 | | 1.2 | -0.51 |
| % 8-10 | -0.7 | 0.61 | 0.27 | -0.09 | -0.48 | 6.78 | 0.04 | -1.5 | -0.15 | 1.5 | -1.4 | 4.9 | | 2.3 | -1.2 | | -0.08 | 2.1 | | 0.27 | -2.6 | | -0.17 | 4.6 | | -0.44 | 4.7 | | -1.7 | 3.1 | | -0.36 | -0.24 |
| DCP |  |  |  |  |  |  |  |  |  |  |  |  | |  |  | |  |  | |  |  | |  |  | |  |  | |  |  | |  |  |
| 2 REP | 0.86 | 6.80 | 0.88 | 4.02 | 0.92 | 6.42 | 0.86 | 7.05 | 0.96 | 4.57 | 0.80 | 11.85 | | 0.85 | 5.97 | | 0.93 | 3.91 | | 0.87 | 6.02 | | 0.92 | 4.38 | | 0.88 | 3.85 | | 0.69 | 3.77 | | 0.84 | 4.65 |
| 4 REP | 0.84 | 7.09 | 0.88 | 4.20 | 0.96 | 4.72 | 0.83 | 7.88 | 0.92 | 6.78 | 0.86 | 10.34 | | 0.73 | 8.08 | | 0.93 | 4.21 | | 0.89 | 5.53 | | 0.94 | 3.90 | | 0.85 | 4.16 | | 0.72 | 3.65 | | 0.82 | 5.17 |
| 6 REP | 0.87 | 6.51 | 0.85 | 4.75 | 0.94 | 5.43 | 0.84 | 7.55 | 0.94 | 5.86 | 0.88 | 9.29 | | 0.69 | 8.00 | | 0.93 | 4.22 | | 0.87 | 6.22 | | 0.94 | 3.77 | | 0.87 | 3.89 | | 0.71 | 3.66 | | 0.83 | 5.08 |
| 8 REP | 0.88 | 6.26 | 0.85 | 4.80 | 0.94 | 5.65 | 0.83 | 7.47 | 0.94 | 6.05 | 0.87 | 9.90 | | 0.70 | 7.74 | | 0.93 | 4.03 | | 0.85 | 6.87 | | 0.92 | 4.37 | | 0.81 | 4.44 | | 0.72 | 3.67 | | 0.81 | 5.29 |
| 10 REP | 0.87 | 6.48 | 0.85 | 4.77 | 0.94 | 5.57 | 0.83 | 7.43 | 0.93 | 6.18 | 0.86 | 10.30 | | 0.71 | 7.76 | | 0.92 | 4.25 | | 0.86 | 6.54 | | 0.92 | 4.20 | | 0.77 | 4.55 | | 0.69 | 3.85 | | 0.82 | 5.10 |
| % 2-4 | -1.7 | 4.1 | -0.79 | 3.6 | 3.7 | -26.4 | -3.1 | 10.5 | -4.7 | 32.6 | 6.3 | -12.8 | | -14.6 | 26.0 | | -0.47 | 6.9 | | 2.0 | -7.1 | | 1.4 | -10.8 | | -2.8 | 6.7 | | 4.2 | -3.3 | | -2.4 | 9.9 |
| % 4-6 | 3.0 | -8.1 | -2.9 | 11.6 | -1.5 | 11.0 | 0.92 | -4.2 | 2.1 | -13.6 | 2.9 | -8.9 | | -4.2 | -0.95 | | -0.08 | 0.39 | | -1.9 | 10.0 | | 0.35 | -3.1 | | 2.0 | -5.9 | | -2.4 | 0.34 | | 1.3 | -1.8 |
| % 6-8 | 1.0 | -3.5 | 0.40 | 1.0 | -0.62 | 3.5 | -1.6 | -1.0 | -0.32 | 2.8 | -1.2 | 5.2 | | 1.3 | -3.3 | | 0.37 | -4.7 | | -2.9 | 9.5 | | -2.1 | 13.6 | | -7.2 | 12.2 | | 1.2 | 0.28 | | -2.5 | 4.1 |
| % 8-10 | -1.1 | 3.2 | 0.04 | -0.71 | 0.18 | -1.3 | 0.07 | -0.57 | -0.28 | 1.9 | -1.3 | 3.4 | | 0.80 | 0.27 | | -1.0 | 5.4 | | 1.6 | -4.8 | | 0.51 | -3.7 | | -3.6 | 2.4 | | -3.8 | 4.6 | | 1.9 | -3.7 |
| TD = typically developing participants; DCP = participants with dyskinetic cerebral palsy. Fl = flexion; Ext = extension; Pro = protraction; Retr = retraction; Rot = rotation, REP = repetitions. | | | | | | | | | | | | | | | | | | | | | | | | | | | | | | | | | |
