## Supplementary table 3 for "Psychometric properties of upper limb kinematics during functional tasks in children and adolescents with dyskinetic cerebral palsy"

| Table S3: Intra-class correlation coefficients (ICC) and standard error of measurement (SEM) and change in ICC and SEM values with each number of different repetitions, expressed in %. | | | | | | | | | | | | |
| --- | --- | --- | --- | --- | --- | --- | --- | --- | --- | --- | --- | --- |
|  | REACH FORWARD | | | | REACH AND GRASP VERTICAL | | | | REACH SIDEWAYS | | | |
|  | Trajectory deviation | | Vmax | | Trajectory deviation | | Vmax | | Trajectory deviation | | Vmax | |
|  | ICC | SEM | ICC | SEM | ICC | SEM | ICC | SEM | ICC | SEM | ICC | SEM |
| TD |  | | | | | | | | | | | |
| 2 REP | 0.85 | 0.04 | 0.85 | 0.11 | 0.60 | 0.02 | 0.90 | 0.08 | 0.58 | 0.04 | 0.71 | 0.15 |
| 4 REP | 0.83 | 0.04 | 0.85 | 0.11 | 0.51 | 0.03 | 0.92 | 0.07 | 0.59 | 0.04 | 0.73 | 0.15 |
| 6 REP | 0.73 | 0.05 | 0.86 | 0.10 | 0.58 | 0.02 | 0.87 | 0.09 | 0.63 | 0.04 | 0.73 | 0.15 |
| 8 REP | 0.75 | 0.04 | 0.86 | 0.10 | 0.59 | 0.03 | 0.87 | 0.09 | 0.61 | 0.04 | 0.76 | 0.14 |
| 10 REP | 0.72 | 0.05 | 0.86 | 0.10 | 0.58 | 0.03 | 0.87 | 0.09 | 0.63 | 0.04 | 0.76 | 0.14 |
| % 2-4 | -1.7 | 2.98 | -14.9 | 0.97 | 1.8 | 11.3 | -0.63 | -5.5 | 1.7 | 4.40 | 2.0 | -1.5 |
| % 4-6 | -12.0 | 20.0 | 11.3 | -6.02 | 6.1 | -7.2 | 1.8 | 15.9 | -4.6 | -1.84 | 0.15 | -0.11 |
| % 6-8 | 1.9 | -3.13 | 1.7 | 0.09 | -2.2 | 3.7 | -0.26 | 0.38 | -0.22 | -0.50 | 4.4 | -5.0 |
| % 8-10 | -3.1 | 3.68 | -0.59 | 2.29 | 1.9 | -1.2 | -0.70 | 0.57 | -0.12 | -2.9 | -0.01 | -3.7 |
| DCP |  | | | | | | | | | | | |
| 2 REP | 0.72 | 0.19 | 0.52 | 0.27 | 0.88 | 0.17 | 0.74 | 0.09 | 0.80 | 0.16 | 0.86 | 0.15 |
| 4 REP | 0.87 | 0.14 | 0.64 | 0.22 | 0.75 | 0.23 | 0.80 | 0.15 | 0.64 | 0.13 | 0.82 | 0.16 |
| 6 REP | 0.85 | 0.17 | 0.70 | 0.20 | 0.75 | 0.23 | 0.82 | 0.14 | 0.70 | 0.12 | 0.80 | 0.17 |
| 8 REP | 0.84 | 0.17 | 0.72 | 0.19 | 0.71 | 0.21 | 0.76 | 0.13 | 0.72 | 0.13 | 0.80 | 0.17 |
| 10 REP | 0.83 | 0.18 | 0.73 | 0.22 | 0.75 | 0.21 | 0.76 | 0.13 | 0.72 | 0.17 | 0.81 | 0.18 |
| % 2-4 | 16.7 | -23.9 | 16.5 | -16.5 | -14.7 | 29.0 | 7.8 | 42.3 | -19.7 | -17.0 | -4.5 | 5.2 |
| % 4-6 | -1.8 | 14.3 | 8.7 | -8.6 | -0.20 | -2.6 | 2.6 | -8.2 | 8.0 | -4.5 | -2.0 | 3.9 |
| % 6-8 | -1.11 | 1.28 | 2.7 | -3.9 | -4.0 | -6.5 | -7.0 | -5.8 | 2.3 | 8.3 | -0.01 | -0.75 |
| % 8-10 | -1.6 | 0.74 | 0.80 | 12.9 | 4.2 | -0.49 | -0.71 | -1.3 | -0.49 | 20.8 | 0.7 | 4.5 |
| TD = typically developing; DCP = dyskinetic cerebral palsy; Vmax = maximal velocity, REP = repetitions. | | | | | | | | | | | | |
