## Supplementary table 4 for "Psychometric properties of upper limb kinematics during functional tasks in children and adolescents with dyskinetic cerebral palsy"

| Table S4: Median and interquartile ranges of standard deviations for joint angles at point of task achievement and spatio-temporal parameters as well as the p-value   for the between-group differences. | | | | | | | | | |
| --- | --- | --- | --- | --- | --- | --- | --- | --- | --- |
|  | **REACH FORWARD** | | | **REACH AND GRASP VERTICAL** | | | **REACH SIDEWAYS** | | |
|  | TD  Median (IQR) | DCP  Median (IQR) | p-value | TD  Median (IQR) | DCP  Median (IQR) | p-value | TD  Median (IQR) | DCP  Median (IQR) | p-value |
| Elevation plane | 2.56 (1.78-3.67) | 3.89 (2.60-6.49) | 0.006* | 2.21 (1.83-4.19) | 3.04 (2.31-7.48) | 0.034* | 3.24 (2.41-4.92 | 4.55 (2.47-7.28 | 0.214 |
| Shoulder elevation | 3.08 (1.43-2.62) | 2.15 (2.37-4.54) | <0.001** | 2.24 (1.70-2.69) | 3.61 (2.11-4.50) | 0.038* | 2.70 (1.69-3.55 | 4.56 (3.37-5.02 | <0.001** |
| Shoulder rotation | 6.35 (3.38-9.73) | 7.42 (4.81-10.66) | 0.330 | 3.70 (2.58-6.49) | 7.65 (4.76-11.12) | 0.088 | 7.51 (6.21-10.51 | 7.79 (5.04-12.33 | 0.687 |
| Elbow flexion/extension | 3.60 (2.44-4.32) | 6.02 (3.59-8.91) | 0.006** | 2.77 (2.03-6.37) | 5.28 (3.01-9.78) | 0.077 | 4.74 (3.43-8.04 | 6.24 (4.72-8.13 | 0.258 |
| Elbow pro/supination | 3.45 (2.29-4.12) | 4.44 (2.92-6.37) | 0.019* | 3.32 (1.97-5.28) | 5.13 (3.34-10.07) | 0.028* | 4.16 (2.98-5.34 | 3.71 (2.66-6.83 | 0.901 |
| Wrist flexion/extension | 3.26 (2.65-4.77) | 6.96 (4.90-19.32) | <0.001** | 5.39 (3.92-7.50) | 6.94 (4.32-5.28) | 0.217 | 3.78 (3.22-4.54 | 8.28 (3.98-12.12 | 0.03** |
| Wrist deviation | 2.98 (2.42-5.16) | 4.84 (3.27-7.51) | 0.012* | 4.00 (2.37-4.71) | 3.73 (2.07-5.58) | 1.000 | 3.28 (2.16-4.10 | 5.73 (3.43-9.67 | 0.004** |
| Scapular pro/retraction | 1.55 (1.10-2.47) | 2.05 (1.43-3.25) | 0.192 | 1.58 (1.10-1.99) | 2.82 (2.06-3.45) | 0.001** | 1.74 (1.34-2.60 | 3.33 (2.55-5.08 | 0.001** |
| Scapular rotation | 1.87 (1.43-2.41) | 2.43 (1.73-3.86) | 0.076 | 1.54 (1.23-2.07) | 3.29 (1.89-5.32) | 0.016* | 2.14 (1.58-2.85 | 3.52 (2.83-6.90 | <0.001** |
| Scapular tilting | 1.82 (1.37-2.41) | 1.99 (1.45-3.05) | 0.231 | 1.72 (1.25-2.10) | 2.53 (1.65-3.83) | 0.016* | 1.76 (1.25-2.26 | 3.20 (2.17-4.04 | <0.001** |
| Trunk flexion/extension | 1.63 (1.09-2.05) | 3.20 (2.05-5.04) | <0.001** | 1.79 (1.07-2.85) | 2.84 (1.92-5.27) | 0.008** | 1.65 (1.23-2.11 | 3.67 (2.58-5.81 | <0.001** |
| Trunk lateral flexion | 1.17 (0.79-2.91) | 3.43 (2.22-5.30) | <0.001** | 1.45 (0.95-2.04) | 2.73 (2.11-4.02) | 0.007** | 1.18 (0.94-2.60 | 3.22 (2.53-4.16 | <0.001** |
| Trunk axial rotation | 2.06 (1.44-2.33) | 3.11 (2.44-5.26) | <0.001** | 2.66 (1.52-3.85) | 3.42 (2.36-5.98) | 0.044* | 2.42 (1.43-3.17 | 3.61 (1.74-6.32 | 0.024* |
| Trajectory deviation | 0.04 (0.03-0.06) | 0.07(0.05-0.14) | 0.001** | 0.02 (0.02-0.03) | 0.08 (0.04-0.22) | <0.001** | 0.04 (0.02-0.05) | 0.09 (0.05-0.10) | <0.001** |
| Maximal velocity (m/s) | 0.09 (0.07-0.12) | 0.12 (0.08-0.18) | 0.076 | 0.09 (0.07-0.12) | 0.12 (0.08-0.16 | 0.010* | 0.13 (0.08-0.15) | 0.14(0.12-0.21) | 0.149 |
| TD = typically developing; DCP = dyskinetic cerebral palsy; IQR = inter-quartile range * = p-value < 0.05; ** = p-value < 0.01 | | | | | | | | | |
