## Supplementary table 1 for "Psychometric properties of upper limb kinematics during functional tasks in children and adolescents with dyskinetic cerebral palsy"

| Table S1: Participant characteristics | | | | |
| --- | --- | --- | --- | --- |
| Participant | **SEX** | **MACS level** | **L/R handed** | **Measured side** |
| DCP1 | M | 3 | R | L |
| DCP2 | F | 1 | R | L |
| DCP3 | F | 1 | R | R |
| DCP4 | F | 2 | L | L |
| DCP5 | M | 3 | L | R |
| DCP6 | M | 2 | R | L |
| DCP7 | M | 2 | R | R |
| DCP8 | M | 2 | L | R |
| DCP9 | F | 2 | R | L |
| DCP10 | M | 3 | R | L |
| DCP11 | M | 3 | R | R |
| DCP12 | M | 2 | R | L |
| DCP13 | M | 2 | R | R |
| DCP14 | F | 3 | R | L |
| DCP15 | F | 3 | R | L |
| DCP16 | M | 2 | L | R |
| DCP17 | F | 2 | R | L |
| DCP18 | M | 2 | L | R |
| DCP19 | M | 2 | R | L |
| DCP20 | F | 3 | R | L |
| TD1 | M | / | R | L |
| TD2 | M | / | R | L |
| TD3 | F | / | R | L |
| TD4 | F | / | R | L |
| TD5 | F | / | L | R |
| TD6 | F | / | R | L |
| TD7 | M | / | R | L |
| TD8 | F | / | R | L |
| TD9 | F | / | R | L |
| TD10 | F | / | R | L |
| TD11 | M | / | L | R |
| TD12 | F | / | R | L |
| TD13 | F | / | R | L |
| TD14 | M | / | R | L |
| TD15 | F | / | R | L |
| TD16 | M | / | R | L |
| TD17 | F | / | L | R |
| TD18 | F | / | R | L |
| TD19 | M | / | R | L |
| TD20 | F | / | R | L |
